# Hypertension control and its relationship with cognitive impairment among adults aged 45 to 80 in China

**DOI:** 10.1101/2023.10.20.23297342

**Authors:** Xin Li, Qian wu, Xing Chen, Yanmin Tang, Beini Fei, Qi zhao, Jing Ding, Xin Wang

**Author notes:** Corresponding to: Jing Ding, MD, Zhongshan Hospital, Fudan University, 180 Fenglin Road, Shanghai 200032, China., Qi Zhao, PhD, School of Public health, Fudan University,138Yixueyuan Road, Shanghai 200032,China. Co-first Author.

## Abstract

This cross-sectional study aimed to investigate the control of hypertension and its association with cognitive impairment in Chinese adults aged 45-80 years. Using cross-sectional surveys conducted in Shanghai and Guizhou from 2019-2021, 9,688 eligible subjects were eventually included. After analyzing the data using statistical methods such as unconditional logistic regression and restricted cubic splines (RCS), we found that severe blood pressure was positively associated with the risk of cognitive impairment. There was no significant association between mild/moderate BP and the risk of cognitive impairment. Moreover, diastolic blood pressure was significantly positively correlated with cognitive impairment. The restricted cubic spline modelresults showed that the associations with cognitive impairment seem to be U-shaped for SBP and linear for DBP. In conclusion, this study shows that uncontrolled hypertension is an independent risk factor for cognitive impairment and that the risk of cognitive impairment increases as diastolic blood pressures continues to rise.

## Introduction

In 2019, the number of people living with dementia worldwide is estimated to be 57.4 million, which is expected to increase to 83.2 million in 2030, 116 million in 2040, and 152.8 million in 2050, dementia is likely to have a critical impact on healthcare systems, care infrastructure and economies. ^1, 2^. According to the statistics of the World Health Organization in 2017, there are more than 10 million people with dementia in China, which is about 21.3% of the total number of people with dementia in the world^3^.

Hypertension is a major risk factor for cardiovascular disease and is the leading cause of disability-adjusted life years globally^4, 5^. Numerous epidemiological studies have demonstrated that as an established risk factor for cerebrovascular disease, emerging evidence suggests that hypertension has an important impact on the development of cognitive impairment, vascular dementia, and Alzheimer’s disease (AD)^6-12^. Since hypertension is a modifiable factor, it is a possible potential mechanism for preventing or delaying age-related cognitive impairment. Thus, the control of pressure levels^13^ seems to moderate the association of hypertension with cognitive decline and dementia, whereas poor blood pressure control may accelerate cognitive decline, but such effects have not yet been sufficiently explored^14-16^.

Although current studies have shown that systolic and diastolic blood pressure is associated with cognitive impairment^17, 18^, most of the previous studies on the association between hypertension and cognitive decline were based on the conventional hypertension definition as BP ≥ 140/90 mmHg^19-24^. The association of borderline hypertension, with BP between 130/80 and 140/90 mmHg, and cognitive decline remains not elucidated^23^. Since the study of borderline hypertension is beneficial to the prevention and control of hypertension, this study investigated the control of hypertension and its relationship with cognitive impairment in middle-aged and older adults aged 45-80 years old in Guizhou and Shanghai, China. In addition, we also investigated the dose-response relationship between systolic and diastolic blood pressure and cognitive impairment, aiming to analyze the thresholds of systolic and diastolic blood pressure for cognitive impairment.

## MATERIALS AND METHODS

### Recruitment of Participants

We selected Shanghai and Guizhou provinces as research sites in mainland China. In Shanghai, we chose representative secondary hospitals and all community hospitals in Qingpu, Xuhui, and Minhang. We chose one representative hospital in each city in Guizhou. From April 2019 to May 2021, we recruited a total of 9,688 research subjects aged ≥45 years and ≤80 years old, conscious, without severe visual, auditory or mental impairment, able to cooperate with the completion of cognitive assessment, and with a clear history of hypertension diagnosis. Participants were excluded if they had:① A clear diagnosis of stroke or other cerebral microvascular diseases; ② Cognitive dysfunction caused by other causes, such as frontotemporal dementia, Lewy body dementia, hydrocephalus, etc.; ③ Past traumatic brain injury, brain tumor, severe nervous system diseases such as central nervous system infection, demyelinating disease, epilepsy, myelopathy; ④ Severe cardiovascular comorbidities and cannot tolerate evaluation; ⑤ Severe visual or hearing impairment, aphasia, mental disorder, gait or balance disorders, etc., who cannot cooperate with cognitive and gait assessment; ⑥ Refuse to participate in the study.

### Assessment of Cognitive Function

Standardized cognitive testing by a trained technician was designed to comprehensively assess cognitive abilities among adults aged 45-80. Global cognitive status was assessed by the mini-cognitive assessment (Mini-Cog). The Mini-Cog is a very brief, widely used cognitive test that includes a memory task and a simplified assessment of the Clock Drawing Test (CDT). We asked subjects to first memorize 3 unrelated words (speak carefully, one second for each item) and immediately repeat these 3 words, and then ask subjects to draw a clock and mark all the numbers, the pointer points to 3:40, and finally, the subjects are asked to recall the 3 words they were asked to remember before.

The subjects’ cognition was scored according to the Mini-Cog scoring standard, which was divided into a short-delayed recall score and clock-drawing test score. The short-delayed recall score was scored according to the number of correctly recalled words, 1 point for each word, for a total of 3 points; The clock drawing test is scored according to the three-point method, in which the outline of the clock is a closed circle and no points are scored, all numbers are correct and the position and order are correct, 1 point, the pointer is correct and points to the correct time, 1 point, a total of 2 points; when the short delay When recalling 3 points, regardless of the score of the clock-drawing test, the cognitive function is considered normal; when the short-latency recall is 1-2 minutes, the cognitive function is normal when the clock-drawing test is 2 minutes, and the clock drawing test is 0-1 minutes. The cognitive function is abnormal; when the delayed recall is 0 points, regardless of the score of the clock drawing test, the cognitive function is considered abnormal.

### Assessment of Blood Pressure

Blood pressure was measured by a professionally trained physician using an electronic monitor (Omron model HEM-7112) and after a 10-minute rest. Diastolic and systolic blood pressure was measured twice at the same time, with the rest between measurements. The average of the two readings was calculated and the blood pressure value was recorded for each participant. In addition, vigorous exercise was not allowed within 30 minutes before measurement. Respondents also reported if a physician had ever told them they had hypertension, and whether they were taking medications for hypertension. We define severe BP: SBP ≥180 mmHg or DBP ≥110 mmHg; moderate BP: SBP of 160–179 mmHg or DBP of 100–109 mmHg; mild BP: SBP of 140–159 mmHg or DBP of 90–99 mmHg; and normal BP: SBP <140 mmHg and DBP <90 mmHg. We categorized SBP and DBP based on a 10-mmHg interval to reflect both the latest ACC/AHA^25^ and the Chinese classification^26, 27^ of blood pressure in adults (SBP: <110,110-119, 120-129, 130-139, 140-149, 150-159, 160-169, ≥170 mmHg; DBP: <60, 60-69, 70-79, 80-89, 90-99, 100-109, and≥110 mmHg)^9^.

### Assessment of Covariates

Covariates were collected by a structured questionnaire; one part is sociodemographic characteristics, including age, sex, and education; the other part is lifestyle habits and comorbidities, including BMI, smoking, drinking, and self-reported hypertension, diabetes, heart disease, arterial disease, hyperuricemia, hyperlipidemia.

### Statistical Analyses

Continuous variables were expressed as the mean ± SD or median (interquartile range), and categorical variables were expressed as frequencies (%). The student’s t-test, ANOVA, and Wilcoxon signed-rank test or Kruskal Wallis test were used to compare the continuous variables; the chi-squared test or Fisher’s exact test was used to compare the categorical variables. The multivariate logistic regression model was used to detect the association between hypertension and cognitive impairment and take the group of normal BP as the referent. When analyzing the relationship between blood pressure values and cognitive impairment, we defined SBP 120 to129 mmHg and DBP 80 to 89 mmHg as the reference group. We use three models: model 1 adjusted age, sex, and city; model 2 adds education level, BMI, smoking, and alcohol consumption; model 3 adds the comorbidities, such as self-reported diabetes, heart disease, and arterial disease, hyperuricemia, hyperlipidemia. For continuous analyses of BP, restricted cubic spline terms with 3 knots were used with reference SBP of 130 mmHg and DBP of 80 mmHg. Analyses were performed using SAS version 9.4 ((Institute Inc., Cary, NC, USA) and R version 4.1.2 (R Foundation for Statistical Computing). A p-value < 0.05 was considered statistically significant and all tests were two-sided.

## Results

### 1. Baseline characteristics

A total of 9,688 participants were included in the analysis. The mean age of the overall population was 66.39±7.19 years. 5,960 (61.5%) were female, 3,728 (38.5%) were male, and the prevalence of cognitive impairment was 49.7% (4,818). Figure 1 shows that the proportion of abnormal blood pressure was 38.03% in the hypertensive population. The blood pressure values of hypertensive people are mainly concentrated in 130 to 139 mmHg (SBP) and 80 to 89 mmHg (DBP). Among hypertensive patients, those with abnormal blood pressure had a significantly higher BMI than those with normal blood pressure and were more likely to have diabetes, dyslipidemia, untreated, and cognitive impairment (Table 1).

**Figure 1.**
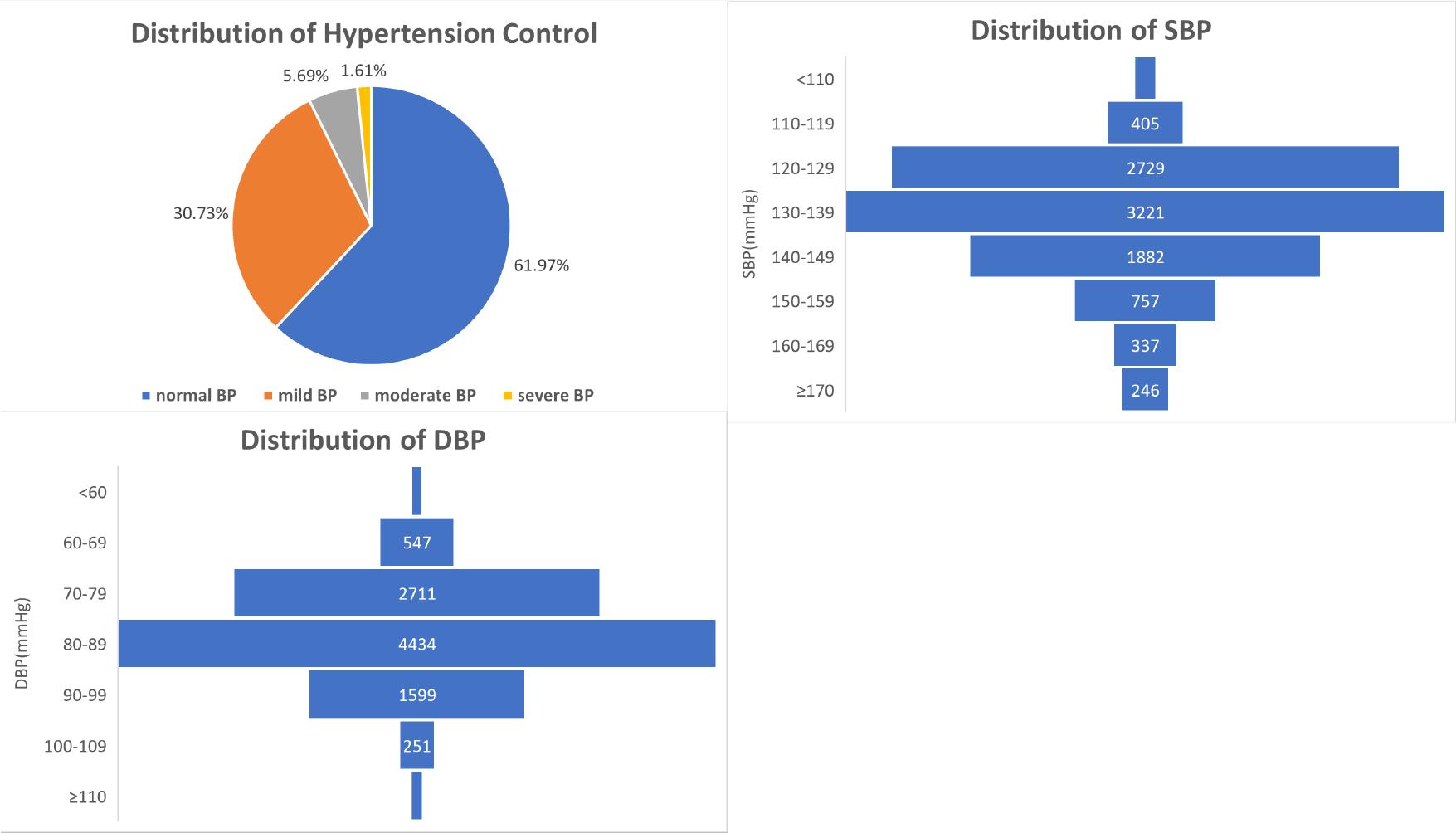
hypertension control and the distribution of SBP/DBP.

**TABLE 1.**
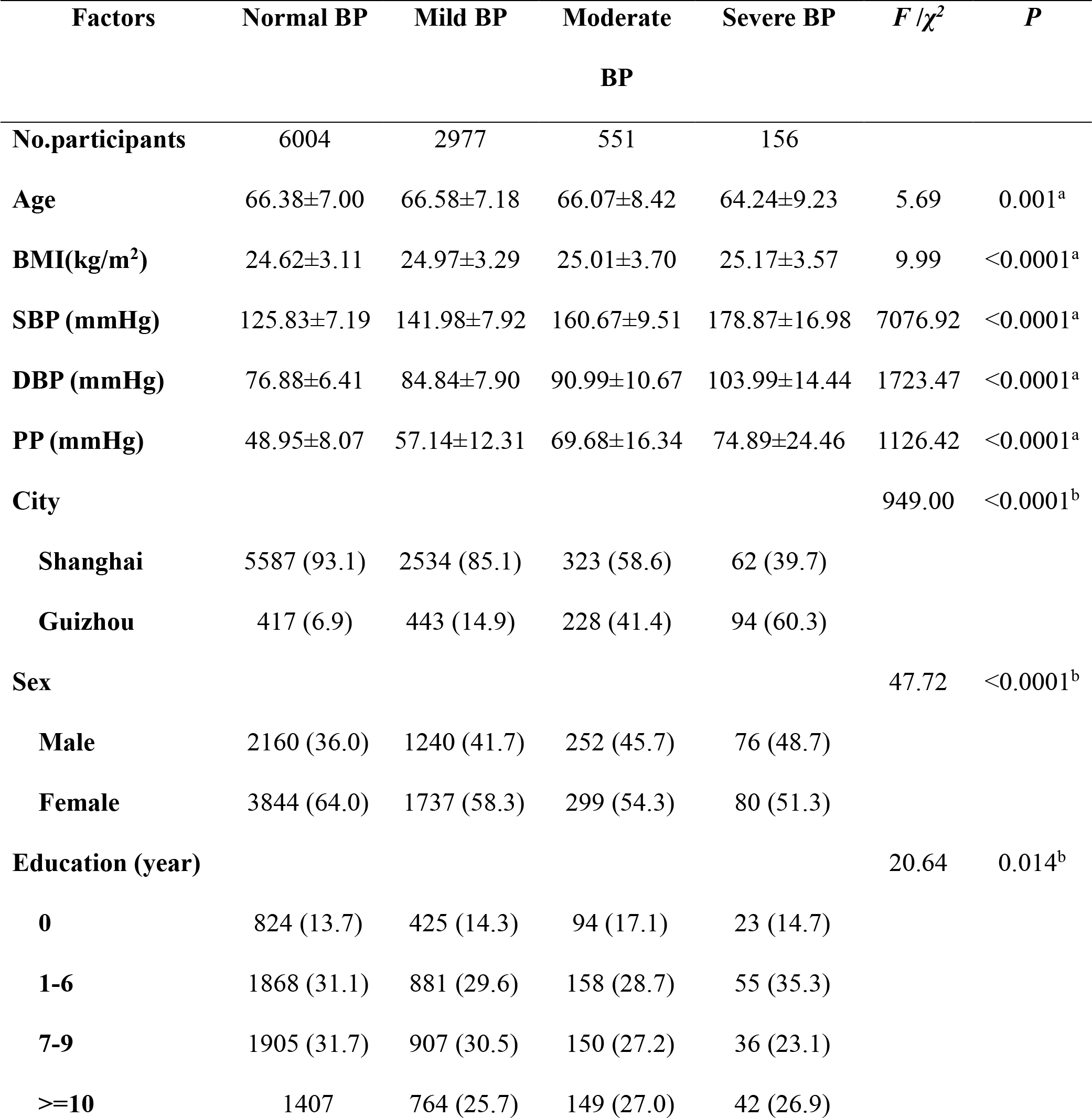
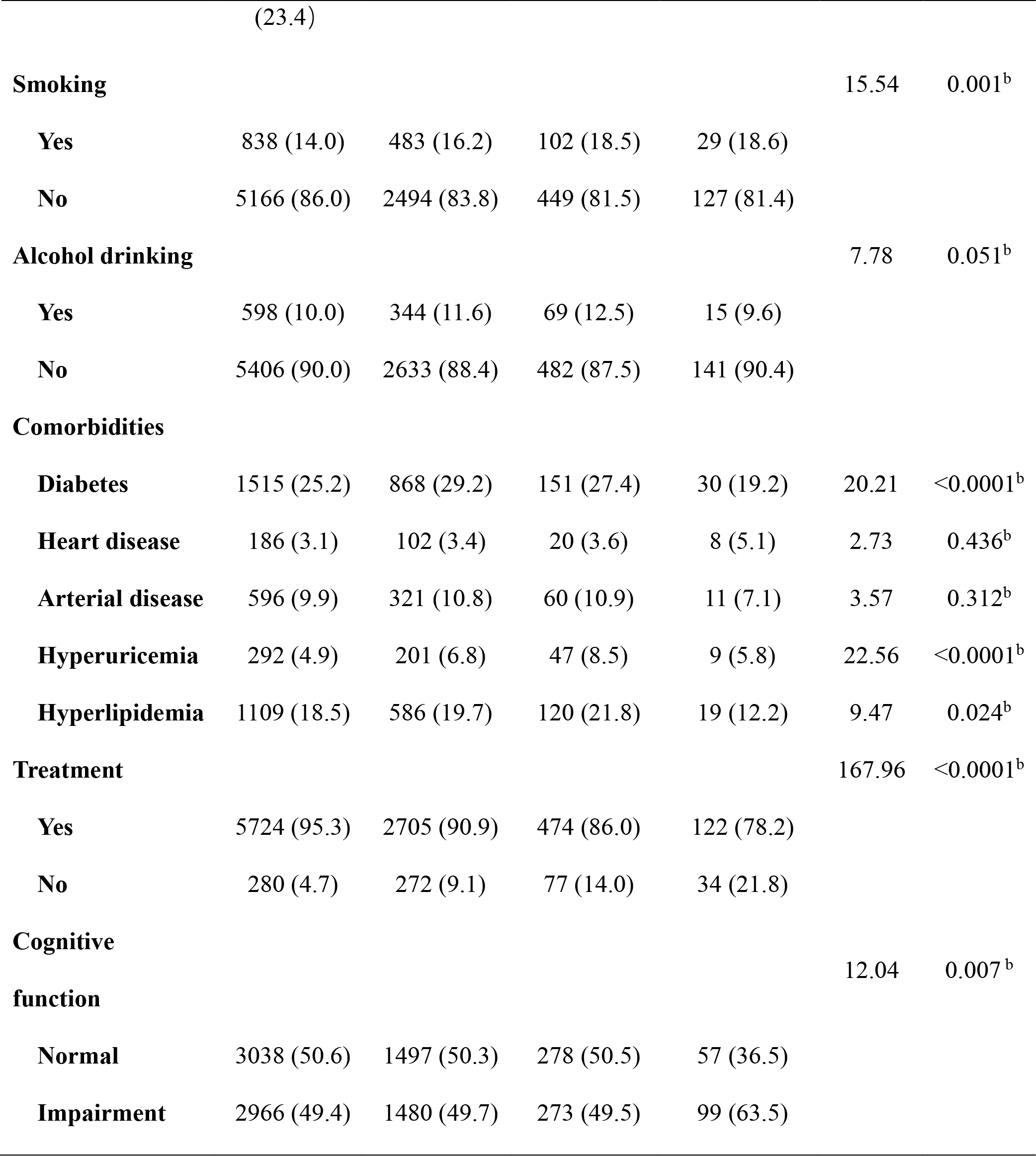
Hypertension and blood pressure of the participants.

### 2. Relationship between blood pressure and the risk of cognitive impairment

We evaluated the association between blood pressure and cognitive impairment with regular logistic regression models for better interpretation (Table 2). The regression model suggested that severe BP was significantly associated with cognitive impairment. Compared with the reference group (normal BP), the adjusted OR (model3) was 1.588 (95%CI 1.105-2.280) for severe BP. And compared with the 81-90 mmHg of DBP, the adjusted OR (model3) was 2.505 (95%CI 1.196-5.249) for >110 mmHg. Each 10-mmHg increase in DBP was associated with a 6.4% increase in the risk of cognitive impairment (95%CI 1.012-1.118). And in the hypertensive patients, the adjusted OR (95% CI) of having cognitive impairment in those with treated hypertension was 0.792 (0.664-0.944) compared with those with untreated hypertensionWe explored the shape of the associations between SBP/DBP and cognitive impairment using the restricted cubic spline curve (Figure 2). There was a nonlinear relationship between the risk of cognitive impairment and SBP in hypertensive participants (P=0.003). And there was a U-shaped relationship between SBP and the risk of cognitive impairment. When systolic blood pressure was lower than 130 mmHg or higher than 155 mmHg, systolic blood pressure was positively associated with the risk of cognitive impairment. And there was a J-shaped for DBP, the OR for cognitive impairment increased as the DBP increased, when DBP was ≥80 mmHg (OR=1.002, 95%CI:1.001-1.003). This suggests that in people with diagnosed hypertension, once SBP or DBP reaches a certain level, the prevalence of cognitive impairment increases further with further increases in SBP or DBP.

**TABLE 2.** Multivariate analysis of the relationship between BP and cognitive impairment.

| MODEL | Model1* | Model2** | Model3*** |
| --- | --- | --- | --- |
| **Hypertension control** |  |  |  |
| **normal BP** | 1(referent) | 1(referent) | 1(referent) |
| **mild BP** | 0.992 (0.907-1.085) | 1.013 (0.921-1.114) | 1.021 (0.928-1.123) |
| **moderate BP** | 0.900 (0.750-1.080) | 0.890 (0.732-1.081) | 0.903 (0.742-1.099) |
| **severe BP** | 1.570 (1.117-2.205) | 1.596 (1.112-2.290) | 1.588 (1.105-2.280) |
| **SBP** |  |  |  |
| **SBP, 10 mmHg** | 0.975 (0.946-1.005) | 0.975 (0.944-1.007) | 0.978 (0.947-1.010) |
| **≤110** | 0.899 (0.695-1.162) | 0.995 (0.758-1.307) | 1.016 (0.773-1.335) |
| **111-120** | 1.152 (1.028-1.290) | 1.115 (0.989-1.258) | 1.104 (0.979-1.246) |
| **121-130** | 1 (referent) | 1 (referent) | 1 (referent) |
| **131-140** | 0.877 (0.786-0.978) | 0.894 (0.796-1.003) | 0.898 (0.800-1.009) |
| **141-150** | 0.954 (0.831-1.094) | 0.948 (0.818-1.097) | 0.953 (0.823-1.104) |
| **151-160** | 1.081 (0.889-1.314) | 1.031 (0.837-1.269) | 1.055 (0.856-1.301) |
| **161-170** | 0.808 (0.605-1.080) | 0.782 (0.573-1.067) | 0.789 (0.578-1.078) |
| **>170** | 1.256 (0.893-1.765) | 1.347 (0.936-1.939) | 1.358 (0.943-1.957) |
| **DBP** |  |  |  |
| **DBP, 10 mmHg** | 1.115 (1.065-1.168) | 1.071 (1.019-1.126) | 1.064 (1.012-1.118) |
| **≤60** | 0.602 (0.457-0.793) | 0.714 (0.532-0.960) | 0.723 (0.538-0.972) |
| **61-70** | 0.866 (0.760-0.985) | 0.935 (0.815-1.073) | 0.946 (0.824-1.086) |
| **71-80** | 0.980 (0.890-1.080) | 0.975 (0.879-1.080) | 0.971 (0.876-1.076) |
| **81-90** | 1 (referent) | 1 (referent) | 1 (referent) |
| **91-100** | 1.150 (0.964-1.373) | 1.078 (0.892-1.303) | 1.076 (0.889-1.301) |
| **101-110** | 0.865 (0.566-1.321) | 0.943 (0.599-1.485) | 0.925 (0.588-1.456) |
| **>110** | 2.896 (1.430-5.864) | 2.540 (1.215-5.313) | 2.505 (1.196-5.249) |
| **Treatment** | 0.786 (0.666-0.926) | 0.782 (0.656-0.932) | 0.792 (0.664-0.944) |

\*Modell: Adjusted age, sex, and city
**\*\*Model2:** Adjusted education level, BMI, smoking, and drinking on the basis of the model1
**\*\*\*Model3:** Adjusted comorbidities on the basis of the model2

**FIGURE 2.**
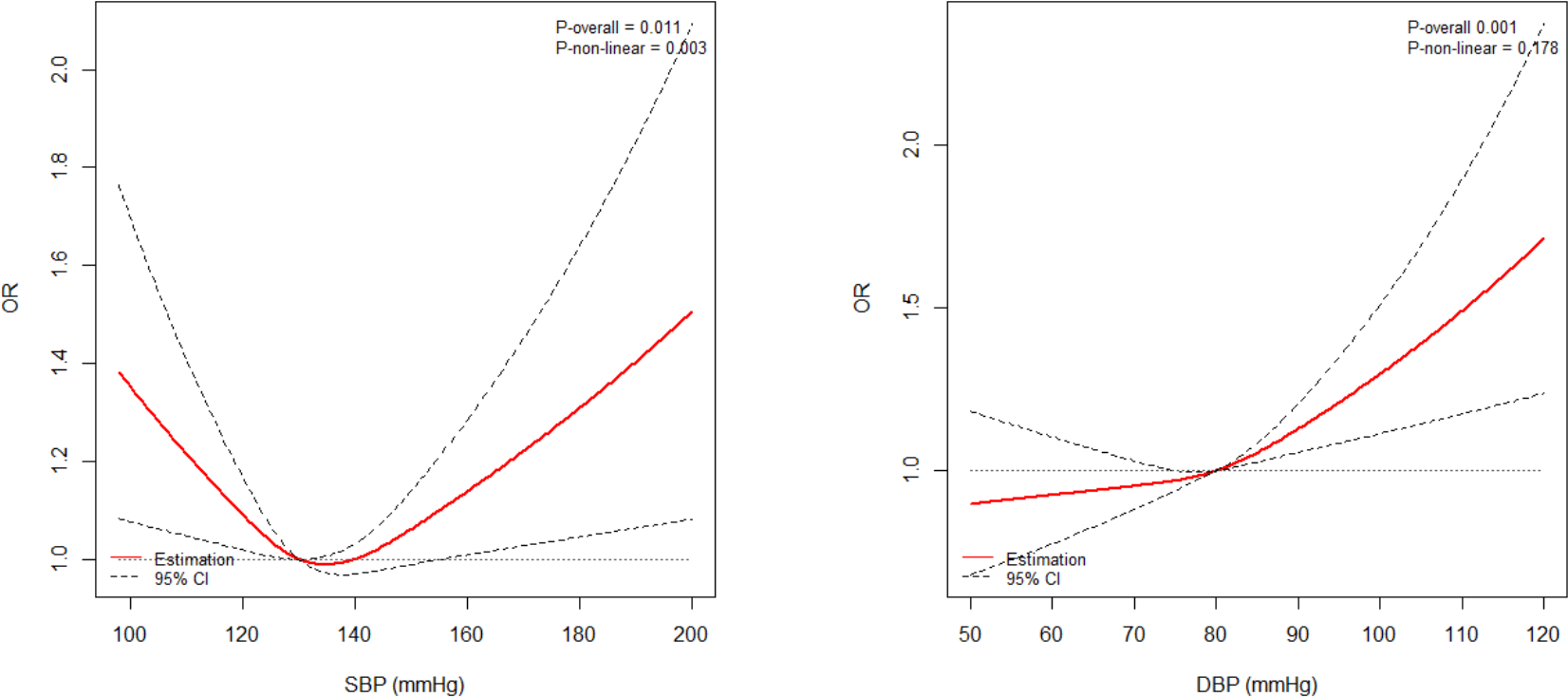
The relationship between SBP/DBP and cognitive impairment (Adjusted cubic spline models showing an association between SBP/DBP and the prevalence of cognitive impairment in hypertensive participants. Models are adjusted for age, sex, city, education level, smoking, alcohol drinking, BMI, diabetes, heart disease, arterial disease, hyperlipidemia, and hyperuricemia. The solid line and long dash line represent the estimated odds ratio and its 95% confidence interval. Knots are at the 25th, 50th, and 75th percentiles.)

## DISCUSSION

Among the middle-aged and elderly people aged 45-80 in China, the control rate of hypertension was 61.97%, which was much higher than the 37.5% reported in other Chinese studies. Although more than one-third of the hypertensive population remains poorly controlled^28^, the significant increase in control rates also indicates that progress appears to have been made in the understanding and control of hypertension over the past decade. However, because the population of this study is mainly Shanghai residents, Shanghai is a developed city in China, and some studies have shown that the hypertension control rate is higher in high-income areas^29^.

In this cross-sectional study in China, we found that in people diagnosed with hypertension, the group with severe BP had a significant impact on cognitive function compared with those with normal blood pressure control. And as diastolic blood pressure gradually rises, and systolic blood pressure is too low and too high, the risk of cognitive impairment increases gradually. Numerous previous epidemiological studies have confirmed that high blood pressure, especially uncontrolled high blood pressure, in the fourth and fifth decade of life is associated with a higher likelihood of cognitive decline 20-30 years later^30-35^.

A prospective study involving 7479 women aged 65-79 in the United States showed that among women with antihypertensive medication, those with BP ≥140/90 mm Hg (uncontrolled BP) were at the highest risk for developing cognitive decline (HR 1.30; 95% CI 1.05, 1.60) compared to women without treatment and BP <140/90 mm Hg (controlled BP). Similarly, in another study using the China Health and Retirement Longitudinal Survey (CHARLS), the same subjects as this study were over 45 years old, and the results showed that uncontrolled hypertension may have a negative impact on 45 years of age P<0.05)^33^. And the results of the ELSA-Brasil study involving 15,105 civil servants (aged 35-74) in Brazil showed that^14^, among treated hypertensive individuals, those with uncontrolled BP showed a more accelerated decline in the memory test and global cognitive score compared with controlled hypertensive individuals. A Chinese study of people over the age of 65 also showed that^36^ among the hypertensive patients, the adjusted OR (95% CI) of having MCI in those with treated hypertension was 0.60 (95%CI 0.42–0.86) compared with those with untreated hypertension, and in those with controlled hypertension was 0.64 (95%CI 0.43–0.93) compared with that non-controlled hypertension (regardless of treatment). Data from the sixth wave of the China Longitudinal Survey of Health and Longevity (CLHLS) conducted in 2011 showed that blood pressure and cognitive function in Chinese elderly over 65 years old also showed a U-shaped relationship. And Mossello et al^37^ also showed that lower daytime systolic blood pressure in 172 older persons with cognitive impairment was associated with a faster cognitive decline. It is suggested that in the hypertensive population, the control of systolic blood pressure needs to be vigilant, and the systolic blood pressure cannot be controlled blindly. A study showed that intensive control of systolic blood pressure significantly reduced incidence of mild cognitive impairment but not possible dementia^38^.The results of a national study involving 19,836 people over the age of 45 in the United States showed^30^ that higher DBP levels were associated with impaired cognitive status. And an increment of 10 mm Hg in DBP was associated with a 7% (95% CI 1.10, 1.14, p = 0.0275) higher odds of cognitive impairment.

Several pathophysiological mechanisms have linked hypertension to cognitive function. Hypertension compromises the structural and functional integrity of the cerebral microcirculation, promoting microvascular rarefaction, cerebromicrovascular endothelial dysfunction, and neurovascular uncoupling, which impair cerebral blood supply^39^. Brain hypoxia can lead to oxidative stress, which in turn affects vascular tone and endothelial function, and induces chronic inflammation^40, 41^. In addition, hypertension can also alter neurochemical transmission, basic cellular functions of neurons, and brain autoregulation, and impairment of these functions can alter the permeability of the blood-brain barrier, allowing toxic substances to enter the brain, and promote neuroinflammation and worsening of amyloidosis^14, 39^. Neuroradiological markers of hypertension-induced cerebral small vessel disease include white matter hyperintensities, lacunar infarcts, and microhemorrhages, all of which are associated with cognitive decline^40^. In addition to this, Petrovic et al^42^ investigated the relationship between midlife hypertension and dementia-related pathology in the Honolulu Asian Study of Aging^42^. They found that systolic blood pressure above 160 mmHg was associated with increased neuritic and neurofibrillary plaque deposition throughout the cortex and hippocampus and decreased total brain volume. In this study, diastolic blood pressure >95 mmHg was associated with increased deposition of neurofibrillary tangles in the hippocampus^42^. The study’s results were limited due to the lack of female participants. José María García-Alberca et al. found that medial temporal atrophy and higher degree of severity of brain white matter hyperintensities among patients with AD and hypertension are associated with the impaired cognitive function^43^.

### Limitations

(1) The mini-cog scale used in this study is a screening scale, which only has a screening effect on cognitive function and cannot be diagnosed; (2) Second, this study only conducted a single test on the subjects of the study. The blood pressure measurement of the patients and the use of antihypertensive drugs and hypertension history are self-reported by the research subjects, so there is a certain error; (3) This study is a cross-sectional study, measuring blood pressure and cognitive status at the same time, it is impossible to infer the two. Therefore, further prospective studies are still needed to verify the effect of blood pressure on cognitive impairment.

### Advantages

(1) This study is a large cross-sectional study based on adults in the community and hospitals, involving people aged 45-80 years old, with large sample size and therefore better representation; (2) This study took into account the gap between the rich and the poor in China. Therefore, both the more developed city Shanghai and the more backward province Guizhou are selected to balance the economic level between regions.

## Conclusions

In conclusion, this study showed that among Chinese adults aged 45-80 years, the empty control rate of hypertension was 61.97%. In this cross-sectional study, severely abnormal blood pressure values were a risk factor for cognitive impairment in a population diagnosed with hypertension, and elevated SBP and DBP were positively associated with an increased risk of cognitive impairment. Therefore, we need to pay attention to the timely treatment of hypertension to control hypertension within a good range, thereby reducing the impact on cognitive function.

## Data Availability

The data that support the findings of this study are available from the corresponding author, upon reasonable request.

## Acknowledgment

We would like to express our gratitude to all the individuals and organizations that contributed to the completion of this research. Without their support and assistance, this study would not have been possible.

## References

1. Estimation of the global prevalence of dementia in 2019 and forecasted prevalence in 2050: An analysis for the global burden of disease study 2019. Lancet Public Health. 2022;7:e105–e125

2. Cataldi R, Sachdev PS, Chowdhary N, Seeher K, Bentvelzen A, Moorthy V, et al. A who blueprint for action to reshape dementia research. Nature Aging. 2023;3:469–471

3. Organization WH. Dementia. 2017

4. Forouzanfar MH, Liu P, Roth GA, Ng M, Biryukov S, Marczak L, et al. Global burden of hypertension and systolic blood pressure of at least 110 to 115 mm hg, 1990-2015. Jama. 2017;317:165–182

5. Zhou B, Perel P, Mensah GA, Ezzati M. Global epidemiology, health burden and effective interventions for elevated blood pressure and hypertension. Nat Rev Cardiol. 2021;18:785–802

6. Qin J, He Z, Wu L, Wang W, Lin Q, Lin Y, et al. Prevalence of mild cognitive impairment in patients with hypertension: A systematic review and meta-analysis. Hypertens Res. 2021;44:1251–1260

7. Shah NS, Vidal JS, Masaki K, Petrovitch H, Ross GW, Tilley C, et al. Midlife blood pressure, plasma β-amyloid, and the risk for alzheimer disease: The honolulu asia aging study. Hypertension. 2012;59:780–786

8. Rönnemaa E, Zethelius B, Lannfelt L, Kilander L. Vascular risk factors and dementia: 40-year follow-up of a population-based cohort. Dement Geriatr Cogn Disord. 2011;31:460–466

9. Yuan JQ, Lv YB, Chen HS, Gao X, Yin ZX, Wang WT, et al. Association between late-life blood pressure and the incidence of cognitive impairment: A community-based prospective cohort study. J Am Med Dir Assoc. 2019;20:177-182.e172

10. Santisteban MM, Iadecola C, Carnevale D. Hypertension, neurovascular dysfunction, and cognitive impairment. Hypertension. 2023;80:22–34

11. She M, Shang S, Hu N, Chen C, Dang L, Gao L, et al. Blood pressure level is associated with changes in plasma aβ(1-40) and aβ(1-42) levels: A cross-sectional study conducted in the suburbs of xi’an, china. Front Aging Neurosci. 2021;13:650679

12. Nordestgaard LT, Christoffersen M, Frikke-Schmidt R. Shared risk factors between dementia and atherosclerotic cardiovascular disease. Int J Mol Sci. 2022;23

13. Köhler S, Baars MA, Spauwen P, Schievink S, Verhey FR, van Boxtel MJ. Temporal evolution of cognitive changes in incident hypertension: Prospective cohort study across the adult age span. Hypertension. 2014;63:245–251

14. de Menezes ST, Giatti L, Brant LCC, Griep RH, Schmidt MI, Duncan BB, et al. Hypertension, prehypertension, and hypertension control: Association with decline in cognitive performance in the elsa-brasil cohort. Hypertension. 2021;77:672–681

15. Messerli FH, Streit S, Grodzicki T. The oldest old: Does hypertension become essential again? Eur Heart J. 2018;39:3144–3146

16. Razquin C, Menéndez-Acebal C, Cervantes S, Martínez-González MA, Vázquez-Ruiz Z, Martínez-González J, et al. Hypertension and changes in cognitive function in a mediterranean population. Nutr Neurosci. 2022;25:612–620

17. Streit S, Poortvliet RKE, Elzen W, Blom JW, Gussekloo J. Systolic blood pressure and cognitive decline in older adults with hypertension. Ann Fam Med. 2019;17:100–107

18. Li C, Zhu Y, Ma Y, Hua R, Zhong B, Xie W. Association of cumulative blood pressure with cognitive decline, dementia, and mortality. J Am Coll Cardiol. 2022;79:1321–1335

19. Yancy CW, Jessup M, Bozkurt B, Butler J, Casey DE, Jr., Colvin MM, et al. 2017 acc/aha/hfsa focused update of the 2013 accf/aha guideline for the management of heart failure: A report of the american college of cardiology/american heart association task force on clinical practice guidelines and the heart failure society of america. Circulation. 2017;136:e137–e161

20. Williams B, Mancia G, Spiering W, Agabiti Rosei E, Azizi M, Burnier M, et al. 2018 esc/esh guidelines for the management of arterial hypertension: The task force for the management of arterial hypertension of the european society of cardiology and the european society of hypertension: The task force for the management of arterial hypertension of the european society of cardiology and the european society of hypertension. J Hypertens. 2018;36:1953–2041

21. Ou YN, Tan CC, Shen XN, Xu W, Hou XH, Dong Q, et al. Blood pressure and risks of cognitive impairment and dementia: A systematic review and meta-analysis of 209 prospective studies. Hypertension. 2020;76:217–225

22. Kivipelto M, Helkala EL, Hänninen T, Laakso MP, Hallikainen M, Alhainen K, et al. Midlife vascular risk factors and late-life mild cognitive impairment: A population-based study. Neurology. 2001;56:1683–1689

23. Ma Y, Hua R, Yang Z, Zhong B, Yan L, Xie W. Different hypertension thresholds and cognitive decline: A pooled analysis of three ageing cohorts. BMC Med. 2021;19:287

24. Knopman DS, Gottesman RF, Sharrett AR, Tapia AL, DavisThomas S, Windham BG, et al. Midlife vascular risk factors and midlife cognitive status in relation to prevalence of mild cognitive impairment and dementia in later life: The atherosclerosis risk in communities study. Alzheimers Dement. 2018;14:1406–1415

25. Whelton PK, Carey RM, Aronow WS, Casey DE, Jr., Collins KJ, Dennison Himmelfarb C, et al. 2017 acc/aha/aapa/abc/acpm/ags/apha/ash/aspc/nma/pcna guideline for the prevention, detection, evaluation, and management of high blood pressure in adults: Executive summary: A report of the american college of cardiology/american heart association task force on clinical practice guidelines. Hypertension. 2018;71:1269–1324

26. [chinese expert consensus on the diagnosis and treatment of hypertension in the elderly(2017)]. Zhonghua Nei Ke Za Zhi. 2017;56:885–893

27. Liu LS. [2010 chinese guidelines for the management of hypertension]. Zhonghua Xin Xue Guan Bing Za Zhi. 2011;39:579–615

28. Lu J, Lu Y, Wang X, Li X, Linderman GC, Wu C, et al. Prevalence, awareness, treatment, and control of hypertension in china: Data from 1·7 million adults in a population-based screening study (china peace million persons project). The Lancet. 2017;390:2549–2558

29. Cheng H, Gu Y, Ma X, Tang H, Liu X. Urban-rural disparities in hypertension prevalence, awareness, treatment, and control among chinese middle-aged and older adults from 2011 to 2015: A repeated cross-sectional study. BMC Cardiovasc Disord. 2022;22:319

30. Tsivgoulis G, Alexandrov AV, Wadley VG, Unverzagt FW, Go RC, Moy CS, et al. Association of higher diastolic blood pressure levels with cognitive impairment. Neurology. 2009;73:589–595

31. Pavlik VN, Hyman DJ, Doody R. Cardiovascular risk factors and cognitive function in adults 30-59 years of age (nhanes iii). Neuroepidemiology. 2005;24:42–50

32. Singh-Manoux A, Marmot M. High blood pressure was associated with cognitive function in middle-age in the whitehall ii study. J Clin Epidemiol. 2005;58:1308–1315

33. Wei J, Yin X, Liu Q, Tan L, Jia C. Association between hypertension and cognitive function: A cross-sectional study in people over 45 years old in china. J Clin Hypertens (Greenwich). 2018;20:1575–1583

34. Liu L, Hayden KM, May NS, Haring B, Liu Z, Henderson VW, et al. Association between blood pressure levels and cognitive impairment in older women: A prospective analysis of the women’s health initiative memory study. Lancet Healthy Longev. 2022;3:e42–e53

35. Naharci MI, Katipoglu B. Relationship between blood pressure index and cognition in older adults. Clin Exp Hypertens. 2021;43:85–90

36. Wu L, He Y, Jiang B, Liu M, Wang J, Yang S, et al. The association between the prevalence, treatment and control of hypertension and the risk of mild cognitive impairment in an elderly urban population in china. Hypertens Res. 2016;39:367–375

37. Mossello E, Pieraccioli M, Nesti N, Bulgaresi M, Lorenzi C, Caleri V, et al. Effects of low blood pressure in cognitively impaired elderly patients treated with antihypertensive drugs. JAMA Intern Med. 2015;175:578–585

38. Rapp SR, Gaussoin SA, Sachs BC, Chelune G, Supiano MA, Lerner AJ, et al. Effects of intensive versus standard blood pressure control on domain-specific cognitive function: A substudy of the sprint randomised controlled trial. Lancet Neurol. 2020;19:899–907

39. Ungvari Z, Toth P, Tarantini S, Prodan CI, Sorond F, Merkely B, et al. Hypertension-induced cognitive impairment: From pathophysiology to public health. Nat Rev Nephrol. 2021;17:639–654

40. Wahidi N, Lerner AJ. Blood pressure control and protection of the aging brain. Neurotherapeutics. 2019;16:569–579

41. Montezano AC, Touyz RM. Molecular mechanisms of hypertension--reactive oxygen species and antioxidants: A basic science update for the clinician. Can J Cardiol. 2012;28:288–295

42. Moonga I, Niccolini F, Wilson H, Pagano G, Politis M. Hypertension is associated with worse cognitive function and hippocampal hypometabolism in alzheimer’s disease. Eur J Neurol. 2017;24:1173–1182

43. García-Alberca JM, Mendoza S, Gris E, Royo JL, Cruz-Gamero JM, García-Casares N. White matter lesions and temporal atrophy are associated with cognitive and neuropsychiatric symptoms in patients with hypertension and alzheimer’s disease. Int J Geriatr Psychiatry. 2020;35:1292–1300

